# Epidemic Situation of Novel Coronavirus Pneumonia in China mainland

**DOI:** 10.1101/2020.02.17.20024034

**Authors:** Youbin Liu, Liming Gong, Baohong Li

## Abstract

**Objective:** Analyze the occurrence of novel coronavirus pneumonia(NCP) in China mainland, explore the epidemiological rules, and evaluate the effect of prevention and control.

**Methods:** From December 1, 2019 to March 4, 2020, Analysis of 80,409 confirmed cases of NCP in China mainland.

**Results:** From December 1, 2019 to March 4, 2020, a total of 80,409 cases of NCP were confirmed in China mainland, a total of 67,466 cases were confirmed in Hubei Province, a total of 49,671 cases were confirmed in Wuhan city. From December 1, 2019 to March 4, 2020, a total of 3,012 cases of NCP deaths in China mainland, the mortality was 3.75% (3012/80,409); A total of 52045 cases of cured in China mainland; The turning point of the epidemic have been reached since February 18.2020 in China mainland; The spread index of NCP gradually declined since January 27. 2020, and the extinction index of NCP rose little by little since January 29, 2020.

**Conclusion:** From December 1, 2019 to March 4, 2020, NCP is under control, and the trend of the epidemic will eventually disappear; The turning point of an epidemic that I’ve created is a great indicator that can calculate the turning date of an outbreak and provide a basis for scientific prevention.

## Introduction

A novel coronavirus was discovered in Wuhan city, Hubei Province on December 1, 2019[1–2]. On February 11, 2020, WHO Director-General announced that the novel coronavirus was officially named “COVID-19(Corona Virus Disease 2019)”, and meanwhile, the international committee for the classification of viruses named “SARS-CoV-2(Severe Acute Respiratory Syndrome Coronavirus 2)”[3], the Chinese Health and Health Commission temporarily named novel coronavirus-infected pneumonia as novel coronavirus pneumonia, abbreviated as“NCP”[4]. The NCP has been included in the B class infectious diseases stipulated in the law of the People’s Republic of China on the prevention and treatment of infectious diseases, and adopted measures for the prevention and control of Class A infectious diseases. According to the “Diagnosis and treatment of pneumonia for novel coronavirus infection” (version1–7), released by the Chinese health commission[5], The incubation period is 1~14 days, generally 3~7 days. Fever, fatigue, dry cough as the main performance[6]. There is clear the route of transmission of respiratory droplet and contact transmission[7], aerosol and the digestive tract of transmission is unclear, the general population susceptibility, the novel coronavirus is sensitive to ultraviolet light and heat, most disinfectants can effectively killed virus, but chlorine has not effectively inactivated virus, should avoid to use hand disinfectant containing chlorine has set. At present, The diagnosed patients were treated with antiviral therapy, serum of the recovered patients, blood purification therapy, immunotherapy, traditional Chinese medicine therapy and a series of symptomatic treatments for each individual. Of course, there are no targeted treatment drugs[8–9]. Because NCP is a new infectious disease, human beings know little about the virus host, species, characteristics of the virus and its epidemiology etc. Therefore, the occurrence, epidemiology and prevention effect of the NCP were analyzed and discussed. It provides a scientific basis for the prevention and treatment of the NCP.

## Data and methods

The data are from the official website of China Health and Health Committee and Hubei Health and Health Committee. From December 1, 2019 to March 4, 2020[10–11]. The standard of diagnosis and treatment for novel coronavirus pneumonia released by the Chinese health commission(version1–7).

The turning point of the epidemic, which is my new creation. the sum of the number of cured patients and the number of dead patients on that day is greater than the number of newly confirmed patients on that day, which is the turning point of the disease. In other words, when the ratio is greater than 1, the epidemic gradually disappears, and when the ratio is less than 1, the epidemic will be continuous.

WPS software was used to analyze and summarize; GraphPad Prism 6 software was used for mapping.

## Results

### 1. Situation of NCP in China mainland

The first case of NCP was reported in Wuhan city of Hubei Province on December 1, 2019. From December 1, 2019 to March 4, 2020, a total of 80,409 cases of NCP were confirmed in China mainland, a total of 67,466 cases were confirmed in Hubei Province, a total of 49,671 cases were confirmed in Wuhan city. It can be seen from Figure 1.A that the cases of the cumulative infection with NCP in China mainland. After treatment, most of the patients have recovered, it can be seen from Figure 1.B that the cases of post-treatment infection with NCP in China mainland. From December 1, 2019 to March 4, 2020, the cumulative deaths with NCP were 3,012 cases, the mortality were 3.75% (3012/80,409). The cumulative cured with NCP were 52045 cases so far, and most of them will be recover.

**Figure 1.**
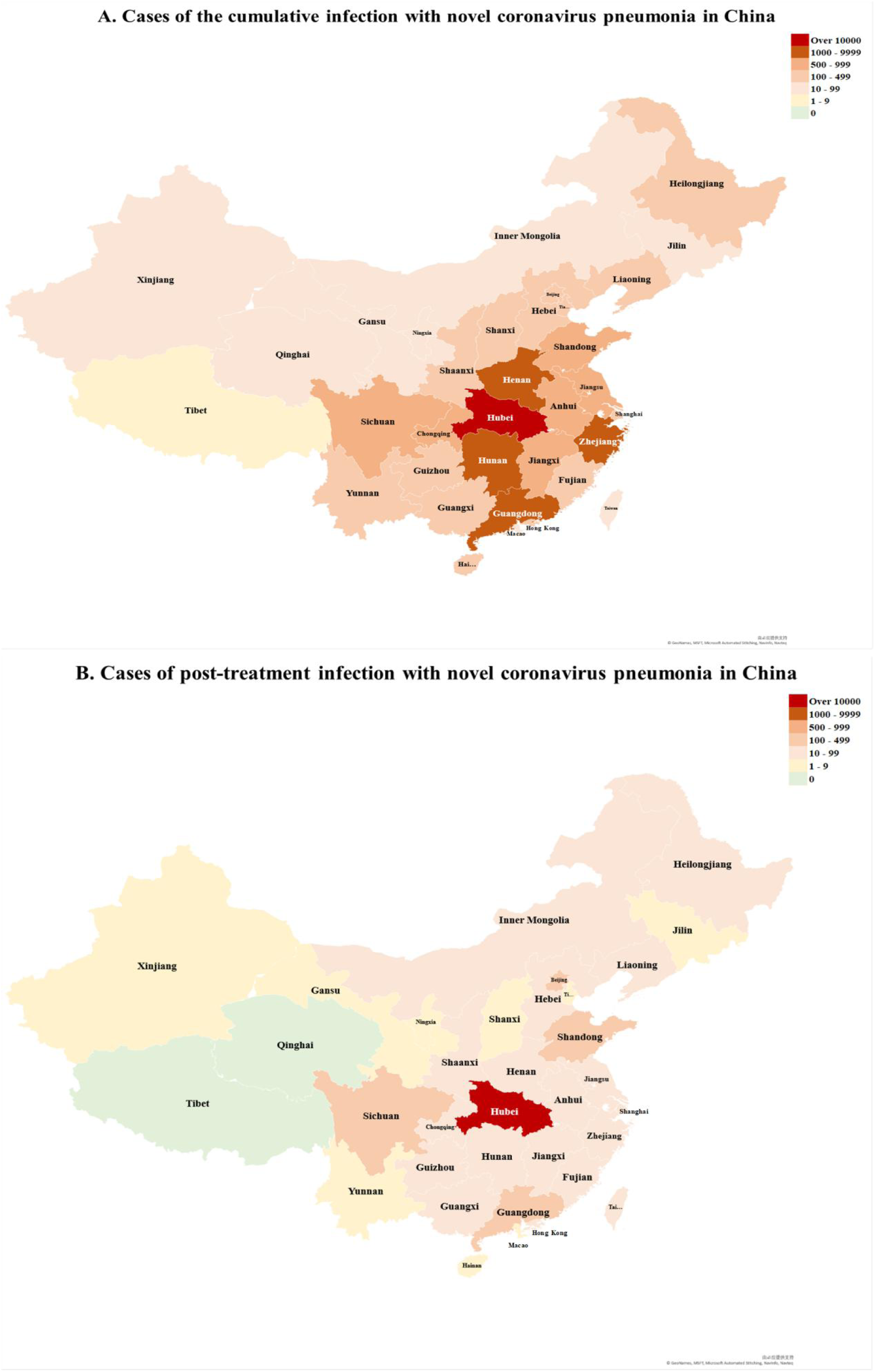
Cumulative cases of NCP in China mainland, More than 10,000 people have been diagnosed with NCP in Hubei province, and more than 1,000 people have been diagnosed with NCP in Guangzhou, Zhejiang, Henan and Hunan(A); After control and treatment, Shandong, Guangzhou, Sichuan and Beijing had residual infections of more than 100 people, while other provinces had fewer than 100 or none (B).

It can be seen from Figure 2 that the number of suspected NCP in China mainland increased by a leap on January 26, 2020. On February 5, 2020, the number of suspected confirmed cases in China mainland reached 5,328. And then it goes down. On February 4, 2020, the number of confirmed cases of NCP in China mainland reached 3887, and since then, the confirmed cases have gradually declined. On February 12, 2020, the number of confirmed cases of NCP in China mainland has exploded to 15152. Since then, the number of newly confirmed cases of NCP in China mainland is continuous decline. From February 3, 2020, the number of newly confirmed cases of NCP is continuous decline in China mainland except Hubei Province, This shows that the control of the transmission of NCP has achieved. Judged from the number of confirmed cases of NCP, controlling the outbreak in China mainland is to control the epidemic in Hubei province, and the key to controlling the outbreak in Hubei province is to control the epidemic in Wuhan city. Eventually, trends suggest that the epidemic will be eliminated.

**Figure 2.**
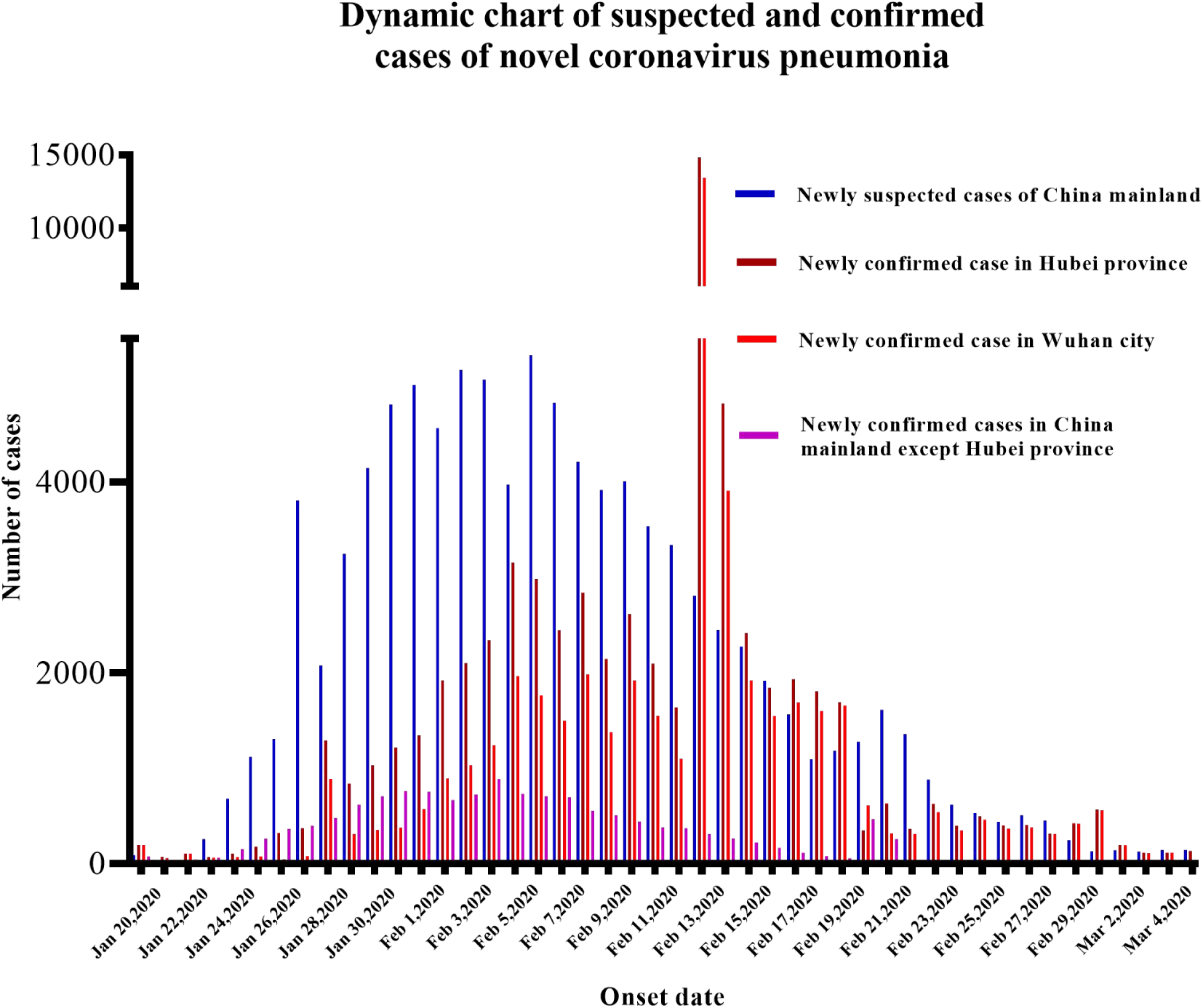
Dynamic chart of suspected and confirmed cases of NCP. On 26 January, 2020, suspected cases of NCP jumped to 3,806; And on 12 February 2020, confirmed cases of NCP exploded to 15,152; On 4 Mar 2020, the number of suspected and confirmed cases is 143 and 139 respectively in China mainland.

### 2. Turning point of the epidemic

The time of the turning point can be seen from figure 4. February 12, 2020 was a turning point of the epidemic in the China mainland except Hubei province, February 18, 2020 was a turning point of the epidemic in the China mainland, and February 19, 2020 was a turning point of the epidemic in Hubei province. From that day on, the total number of cured and dead patients was greater than the number of new patients.

**Figure 3.**
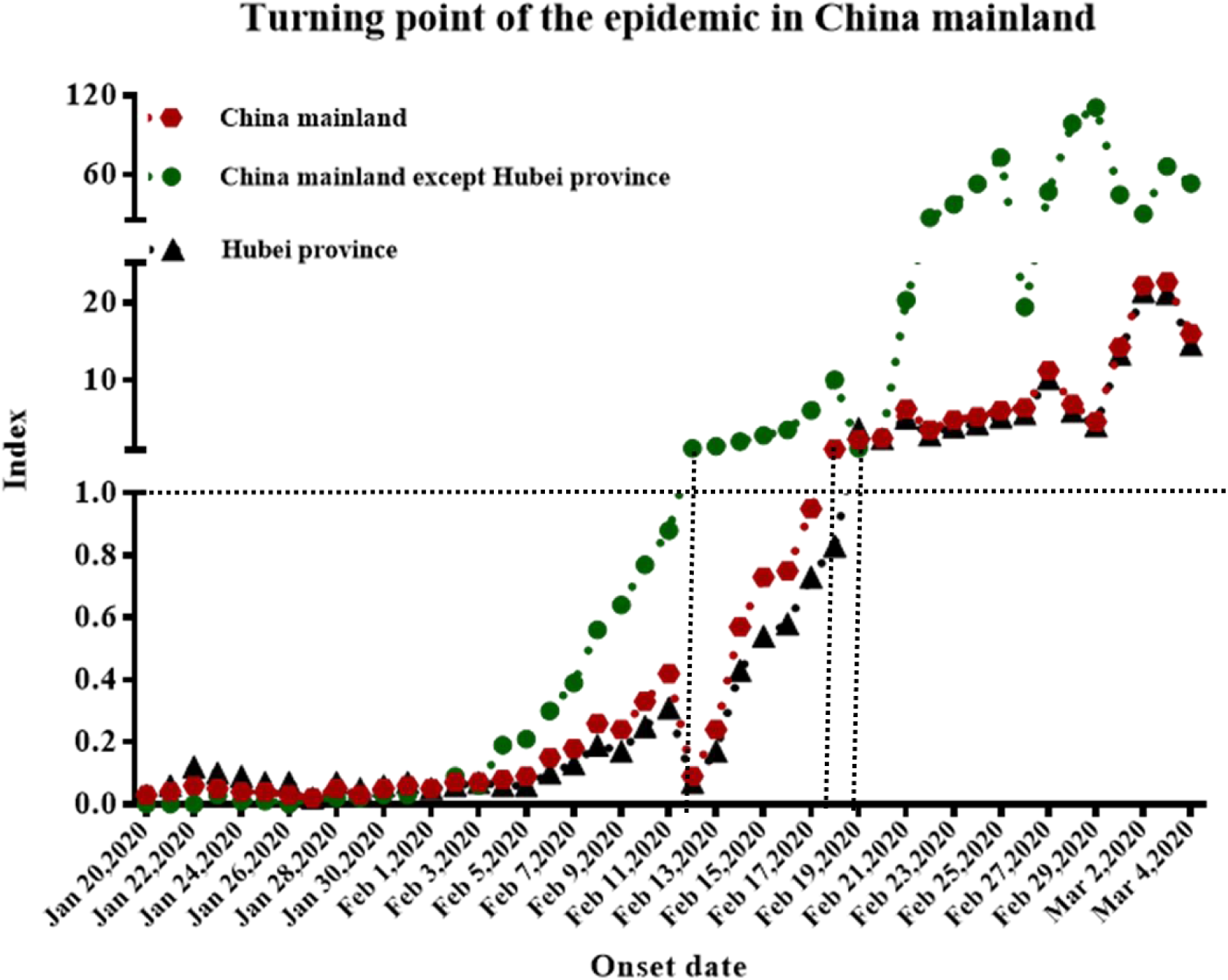
Turning point of the epidemic in China mainland. The date of turning point is February 12 in China mainland except Hubei province; The date of turning point is February 18 in China mainland. The date of turning point is February 19 in Hubei province.

### 3. Epidemic spread index and extinction index

The spread index refers to the ratio of the number of newly confirmed cases on the day to the number of existing cases on the previous day. This index equal to 0, which means that the epidemic does not spread and there are no new cases; The smaller the spread index, the slower the spread of the epidemic; the larger the spread index, the faster the spread of the epidemic. From the Figure 4.A, It can be seen that the spread index of the NCP in China mainland has gradually decreased since January 27, 2020, indicating that the spread rate of the NCP is slowing down. It can be seen the detain from the Figure 4.A.B.C.

The extinction index is the ratio of the number of people cured and death that day to the number of existing cases the previous days. When the extinction index is larger, the epidemic dies faster. When the extinction index is greater than spread index, the epidemic gradually disappears; And when the extinction index is less than spread index; the epidemic will be go on for some time. The ratio of the extinction index to the extinction index is equal to the turning point. The detain can be seen from figure 4.A.B.C.

**Figure 4.**
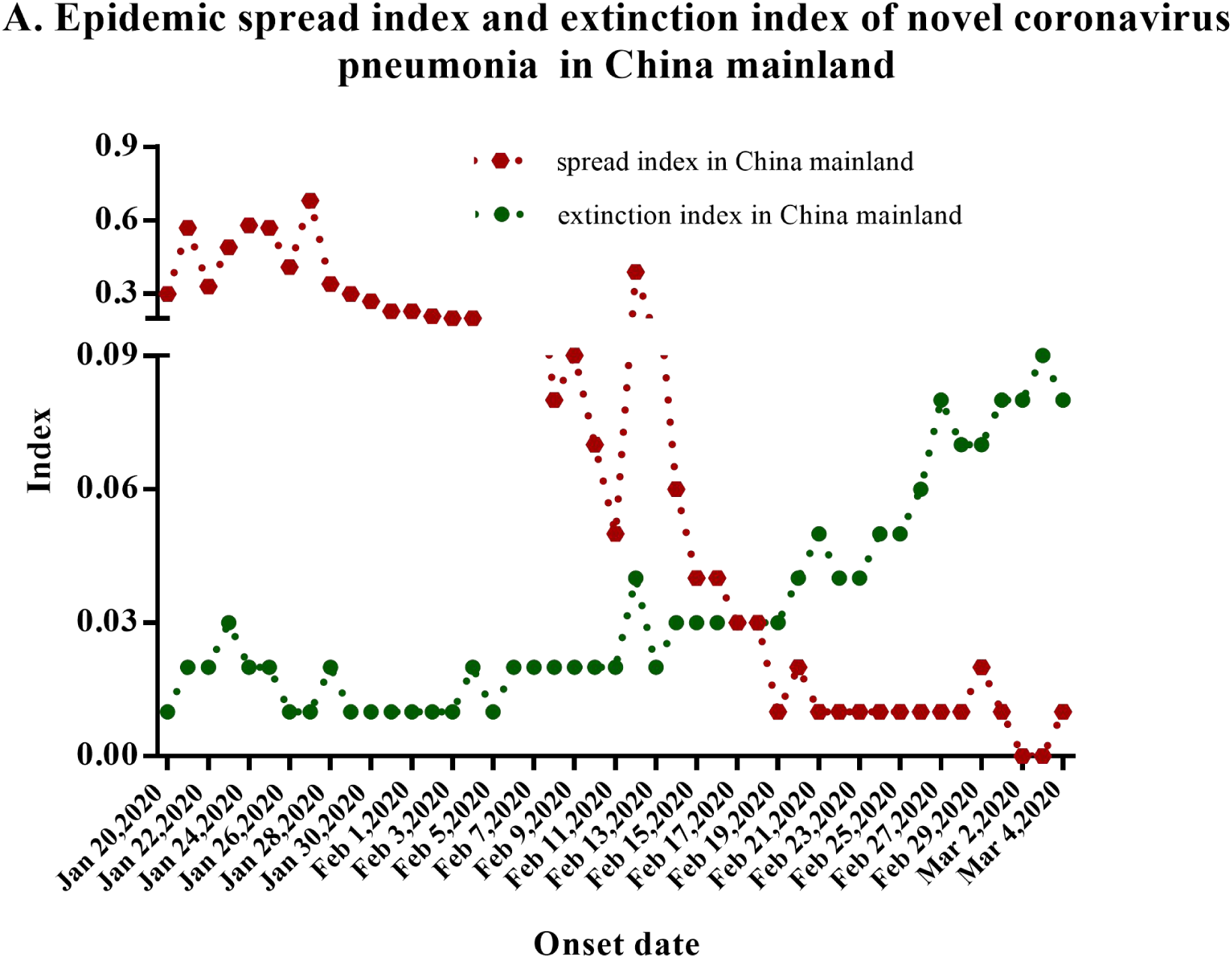
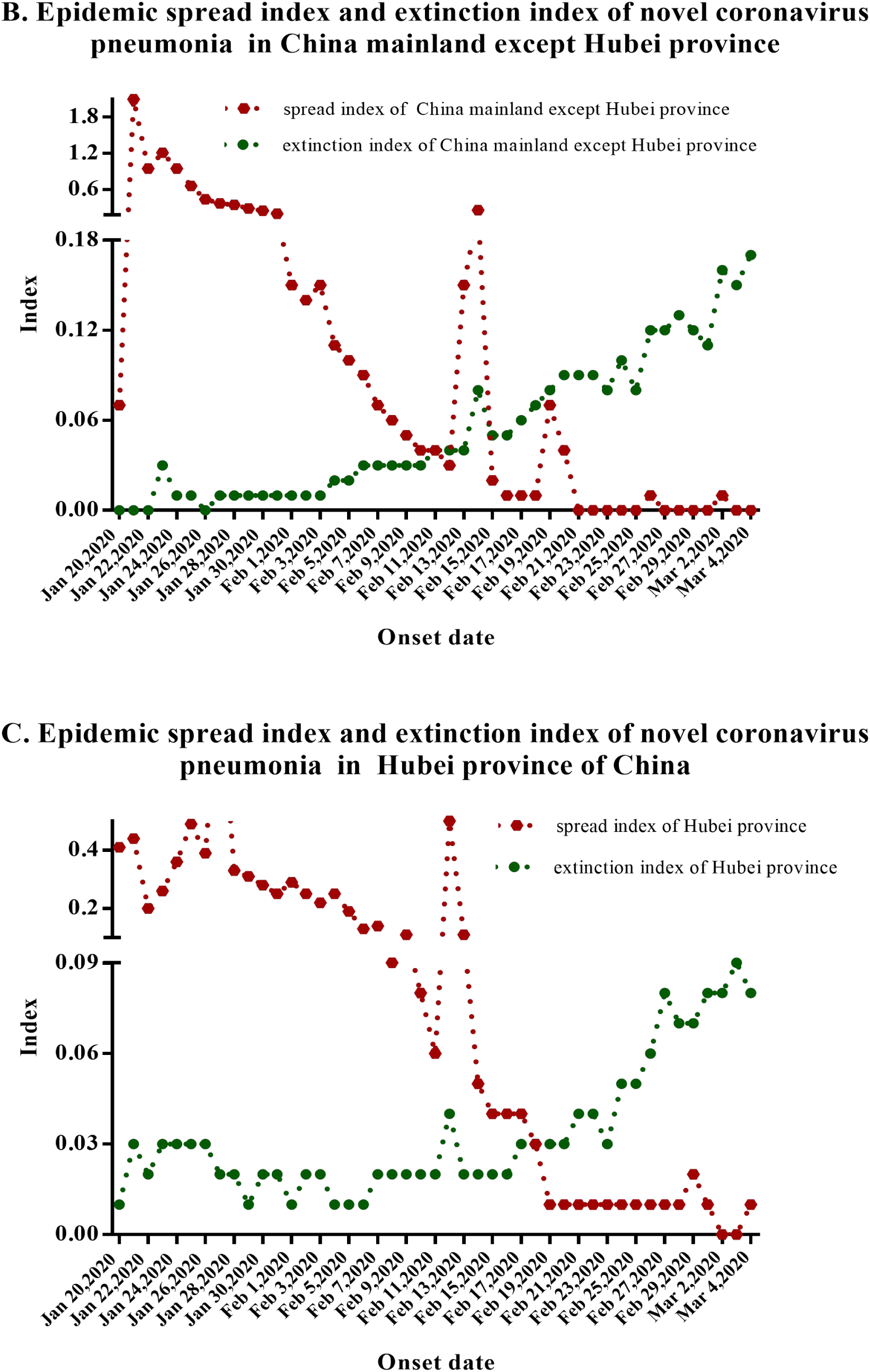
Epidemic spread index and extinction index of NCP in China mainland. The spread index decreases gradually and the extinction index increases gradually. However, as can be seen from figure A, the extinction index of China mainland is still low. At the same time, it can be seen from figure B that in mainland China except Hubei province, from February 12, 2020, the extinction index is greater than 1, indicating the turning point of the NCP.

## Discussion

According to the “Epidemic update and risk assessment of 2019 Novel Coronavirus” released by the Chinese center for disease control and prevention on January 28, 2020[12], the 2019 novel coronavirus is currently believed to be from wild animals, but the details are unclear. South China agricultural university, found beta coronavirus in the pangolin, it is need to further research whether it belong to novel coronavirus or not. South China seafood wholesale market is the main source of infection in Wuhan city of Hubei province. The early cases of NCP were mainly imported infections directly or indirectly from Wuhan[13]. According to the analysis of the current infection situation, Hubei province, is a NCP infections most populous province, followed by Guangzhou province and Zhejiang province, which is related to the economic development and intensive personnel exchanges; Henan province and Hunan provinces also had higher cases of NCP, which is related to neighboring of Hubei province. On January 26, 2020, the suspected cases of the NCP showed a rapid increase. According to the information that the general incubation period of the disease is 3–7 days and the diagnosis needs 2 days, the rapid increase of the suspected cases may be related to the banquet held by the community of Baibuting community in Wuhan city on January 18, 2020. On February 4, 2020, the confirmed cases of NCP has reached 38872. On February, 2020, the suspected cases of NCP has reached 5328. since then, suspected cases and confirmed cases were gradual decline, It may be related to the traffic control in Wuhan, as well as the first-level response to major public health emergencies and relevant control measures launched by several provinces on January 23, 2020, But during the Spring Festival, a time of traditional Chinese holiday, more than 10 million people in the city of Wuhan, the source of the disease, have made it difficult to control the disease because of the movement of people. The gold standard for diagnosing pneumonia caused by novel coronavirus infection is nucleic acid detection[5]. However, the novel coronavirus mainly infects the lower respiratory tract and is difficult to sample. At same time, the kit has high specificity, low sensitivity and long period, which makes difficult to timely diagnosis. After the recommendation of related experts, patients with imaging characteristics of NCP were considered as the standard of clinical diagnosis in Hubei province. This is also one of the important reasons for the explosive increase in the confirmed cases in Hubei on February 12,2020. those patients who had negative nucleic acid test in the early stage, but had CT image features of the NCP[5,14]. After that, screening and diagnosis of novel coronaviruses have been accelerated.

The Chinese government took a series of effective measures, including extending holidays, stopping large gatherings and events, implementing traffic control and grid management, to prevent the further spread of the disease. With the establishment of Huoshenshan Hospital, Leishenshan Hospital and Cabin Hospital, designated hotels were set up to isolate suspected cases of the NCP, and Doctors and nurses from all over the country retrograde to Hubei province to diagnose and treat the patients. The number of people cured of the NCP and the cure rate increased gradually. The number of deaths and mortality of NCP decreased day by day. The reported case mortality of the NCP is lower than that of SARS and MERS [15–16], and the death cases were mainly middle-aged and elderly[1]. The basic reproduction number(R0) is an index that reflects the ability of the virus to transmit[17].The basic reproduction number(R0) is defined as the number of second-generation cases that can be infected under ideal conditions when a case enters a susceptible population. On January 23, 2020, the WHO estimated that the basic reproduction number of the novel coronavirus was 1.4-2.5[18]. The infectious power of the novel coronavirus was lower than that of SARS and MERS [19–22].

As for the “turning point” of the NCP, The ratio of the number of patients cured and the number of patients who died that day to the number of newly diagnosed patients that day is more sensitive. This is a good indicator of turning points. This indicator relates to the lethality and transmissibility of the virus, as well as to the use of artificial blockade and treatment. It’s also equal to the ratio of the extinction index to the spread index. The spread index reflects the ability of the epidemic to spread, and the spread index of the novel coronavirus is not high. Beginning on January 27, 2020, the spread index gradually decreased in China mainland. To some extent, the turning point in controlling the spread of NCP appears, but the onset of the novel coronavirus has a confidentiality and the novel coronavirus can also be infected during incubation period, at the same time, many people return back to work after the Spring Festival and the cumulative cases of NCP is still more, All these factors lead to that the NCP was exist for a period of time. Extinction index reflects the extinction rate of the disease. Because there is no specific drug against the NCP and the course is long and the cumulative number of infections is large. At last, the turning point of the epidemic appeared in China mainland since February 18, 2020; The spread index of NCP gradually decreases and the extinction index gradually increases. After the turning point, the extinction index is greater than the spread index, indicating that the NCP is under control and eliminate in the end in China mainland.

Recently, novel coronavirus outbreaks occurred in other parts of the world. We hope those countries in the world can help each other, learn from each other. From the point of view of epidemic, We should work in three aspects: controlling the source of infection, cutting off the route of transmission, and preventing susceptible people, and the novel coronavirus will soon be eliminated in the world. At last, special thanks to all individuals and countries for their unselfish attention and assistance for China.

## Data Availability

All data is public, which came from the official website of China Health and Health Committee and Hubei Health and Health Committee.

http://www.nhc.gov.cn/xcs/yqtb/list_gzbd.shtml

http://www.nhc.gov.cn/xcs/yqtb/list_gzbd.shtml

